# Enhancing Chemotherapy Response Prediction via Matched Colorectal Tumor-Organoid Gene Expression Analysis and Network-Based Biomarker Selection

**DOI:** 10.1101/2024.01.24.24301749

**Authors:** Wei Zhang, Chao Wu, Hanchen Huang, Paulina Bleu, Wini Zambare, Janet Alvarez, Lily Wang, Philip B. Paty, Paul B. Romesser, J. Joshua Smith, X. Steven Chen

**Author notes:** Correspondence and requests for materials should be addressed to J.J.S., X.S.C.

## Abstract

Colorectal cancer (CRC) poses significant challenges in chemotherapy response prediction due to its molecular heterogeneity. This study introduces an innovative methodology that leverages gene expression data generated from matched colorectal tumor and organoid samples to enhance prediction accuracy. By applying Consensus Weighted Gene Co-expression Network Analysis (WGCNA) across multiple datasets, we identify critical gene modules and hub genes that correlate with patient responses, particularly to 5-fluorouracil (5-FU). This integrative approach advances precision medicine by refining chemotherapy regimen selection based on individual tumor profiles. Our predictive model demonstrates superior accuracy over traditional methods on independent datasets, illustrating significant potential in addressing the complexities of high-dimensional genomic data for cancer biomarker research.

## Introduction

Colon and rectal cancer remains a major global health challenge [1], ranking as the third most prevalent cancer diagnosis and the third leading cause of cancer-related mortality for both men and women in the United States. It is estimated that more than 150,000 new cases will be diagnosed in the coming year, with >50,000 of those being rectal cancer (RC). It is expected that 52,000 deaths are expected to occur relative to both colon and RC this year [2]. Chemotherapy remains central to the treatment of both colon and rectal cancers and centers around drugs such as 5-fluorouracil (5-FU), Oxaliplatin, and Irinotecan. However, the effectiveness of these treatments varies considerably due to the molecular heterogeneity of colon and rectal tumors, with response rates ranging widely. For instance, the use of 5-fluorouracil (5-FU) in combination with leucovorin as a first-line treatment for metastatic disease shows an overall response rate of approximately 20-30%. Moreover, the introduction of irinotecan or oxaliplatin to this regimen increases the response rate to about 40-50%, demonstrating significant variability in treatment efficacy [3]. Further, this variability is specifically seen in RC where the response to triplet chemotherapy (e.g., FOLFIRI (5-FU, Oxaliplatin, and Irinotecan)) is associated with better prognosis than standard therapy [4, 5]. In addition, given the fact that some RC patients may only require upfront chemotherapy for cure, the proper selection of neoadjuvant chemotherapy will become even more critical [6]. Variability in treatment response highlights the critical need for precise predictive models to better forecast individual chemotherapy responses. Accurate prediction models are essential not just for enhancing treatment effectiveness, but also for avoiding unnecessary side effects in non-responsive patients, ultimately leading to more personalized and effective cancer care.

Current commercially available colorectal cancer gene signature panels, like OncotypeDX, ColoPrint, and others, primarily serve prognostic purposes, but their effectiveness in predicting neoadjuvant therapy response is not well-established [7–12]. Moreover, the likelihood of individual cancer biomarkers reaching clinical significance remains notably low, a reality shaped by multiple factors. One notable challenge is that prediction models derived from cell lines often fail in human tumors [13, 14]. In addition, intratumoral heterogeneity (ITH) via stromal cells in the tumor microenvironment may hide subtle gene expression alterations associated with genetic diversity and heterogeneity within the tumor epithelium [15]. Finally, the complexity of genomic features (i.e., Curse of dimensionality) presents a key challenge to prediction tasks involving the drug response [16–18]. This means the top genomic features selected from primary tumors may not be predictive in different patient cohorts partially due to the false positive signals from high-dimensional molecular data, which was shown in the colorectal cancer biomarker discovery [7–10].

Tumor organoids, three-dimensional cell culture systems, offer a revolutionary approach in cancer research. They closely mimic the structural and functional attributes of original tissues or organs, providing a more accurate representation of human tumors. These organoids preserve the genetic, phenotypic, and behavioral characteristics of their source tumors, making them highly relevant for drug discovery, treatment response tracking, and personalized medicine. Their ability to maintain the heterogeneity of the source tumors enhances the study of tumorigenesis, drug screening, and precision medicine, presenting a significant advantage over traditional two-dimensional cell cultures. Previously, our group has shown that RC organoids correspond to the patient-specific outcomes observed including chemotherapy response and clinical outcomes [19]. This advancement facilitates more reliable investigations into tumor pathogenesis, offering a promising platform for cancer research and clinical applications [20, 21].

Recent advancements in colon and RC organoid research demonstrate effective replication of the intricate cellular diversity and molecular heterogeneity seen in patient tumors, thus providing an essential tool for exploring disease development, progression, and treatment responses [19, 22–25]. The transcriptome data from organoids has been used to predict anti-cancer drug efficacy [26]. Moreover, harnessing the power of colon and RC organoid models and integrating molecular data from matched primary tumors and organoids allows an opportunity to identify novel biomarkers that predict treatment response. Directly selecting markers from tumor samples can result in a high rate of false-positive signals due to the vast number of features relative to the number of observations, which can obscure true biological signals. To mitigate this issue, organoids serve as an amplified biological system that retains the complexity of the original tumor but in a more controlled environment that allows for clearer observation of treatment responses. By comparing the molecular profiles of organoids with those of the corresponding tumor samples, we can more effectively filter out noise and retain robust signals that reflect intrinsic tumor biology. A previous study showed that a cancer-cell intrinsic gene expression signature has excellent predictive performance by minimizing intratumoral heterogeneity bias in colon and RC prognostic/predictive classification [27]. Another recent study found that tumor intrinsic immune signatures developed via a matched organoid–primary tumor system are effective tissue biomarkers of prognosis in colon and RC [28], indicating organoids reflect intrinsic properties of the tumor and its microenvironment [19, 29–33].

Here, we developed new strategies that leverage matched colorectal tumor and organoid transcriptome data and the consensus gene network approach to identify key gene expression biomarkers predictive of 5-FU-based chemotherapy response. Our results indicate that these tumor-based biomarkers are strongly correlated with patient survival outcomes when treated with specific chemotherapeutic agents. This innovative approach marks a notable shift towards precision medicine, offering the potential to customize therapeutic strategies based on individual colorectal patient profiles to enhance treatment efficacy.

## Results

### An Overview of the Integrative Analysis of Matched Colorectal Tumor and Organoid Data for Chemotherapy Response Prediction

This study employs a unique integrative analysis approach, as depicted in **Figure 1**, focusing on matched colorectal cancer (CRC) tumor tissues and patient-derived organoids. The analysis begins by leveraging gene expression data from both CRC tumors and corresponding organoids, drawn primarily from datasets GSE171680 and GSE171681, which include a substantial number of matched samples (87 samples)[28]. Given the lack of direct drug response data in these matched datasets, we next integrated an additional colorectal cancer organoid dataset, GSE64392, which contained IC50 values. The detailed information of three training datasets is listed in **Table 1A**.

**Figure 1.**
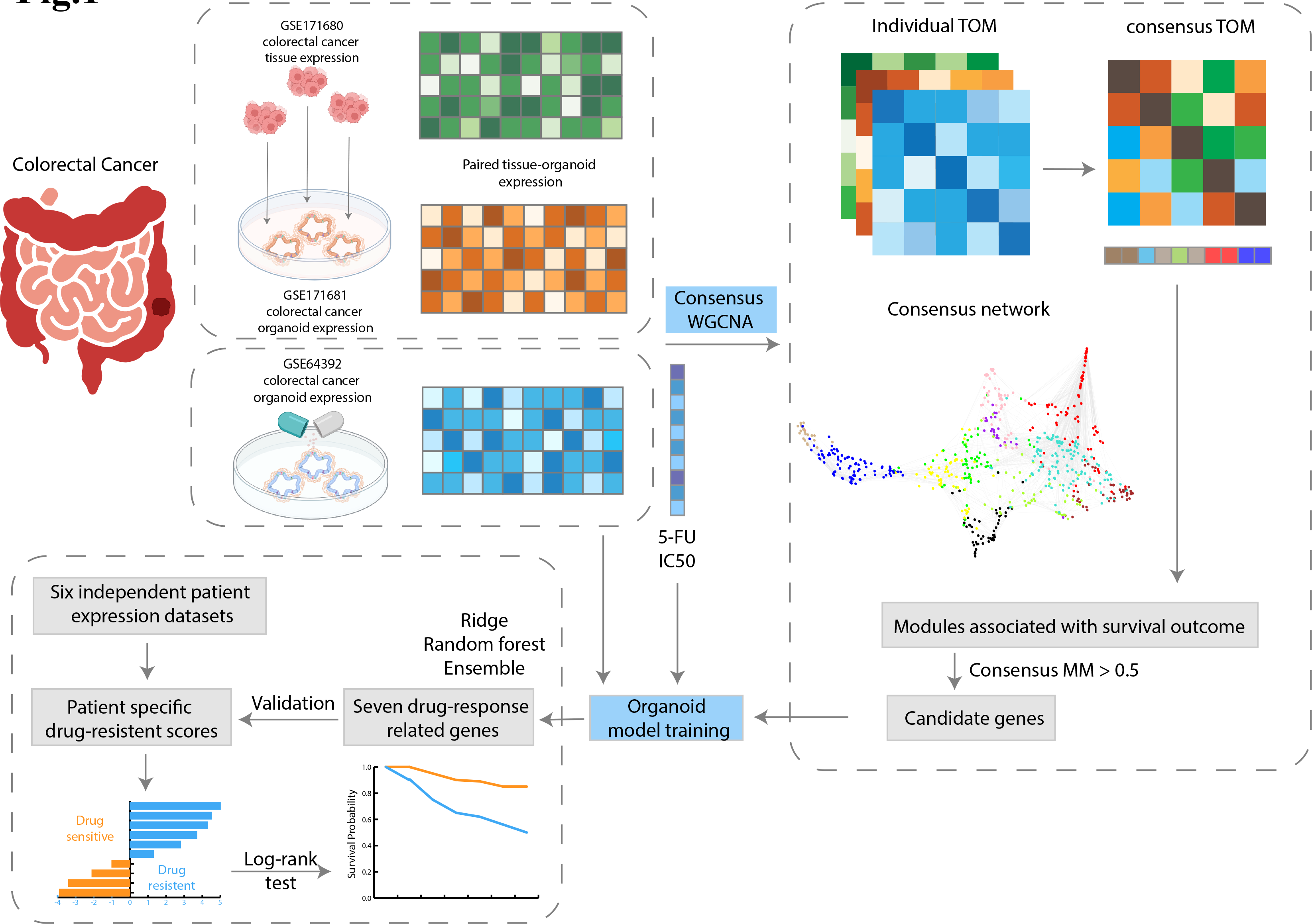
Workflow for the idenfication of gene biomarkers with WGCNA and development of organoid model. The process iniWates with the selecWon of three colorectal cancer datasets for a consensus WGCNA. PotenWal gene biomarkers are then chosen to train the organoid models with GSE64392 and its drug response 5-FU. Subsequently, genes related to drug response are identified and uWlized for prognosis tesWng on six independent colorectal cancer paWent expression datasets.

**Table 1A.** Gene expression training datasets Table 1B Gene expression testing datasets.

| Dataset | Data type | Array type | Number of genes | Number of samples | Have drug response | Number of OS events | Median OS follow-up time | Number of RFS events | Median RFS follow-up time |
| --- | --- | --- | --- | --- | --- | --- | --- | --- | --- |
| GSE171680 | Tissue | RNAseq | 20501 | 87 | No | 15 | 38.27 | 19 | 29.90 |
| GSE171681 | Organoid | RNAseq | 20501 | 87 | No | 15 | 38.27 | 19 | 29.90 |
| GSE64392 | Organoid | Microarray | 25988 | 19 | Yes | NA | NA | NA | NA |

**Table 1B.** Gene expression testing datasets.

| Datasets | Array type | Number of genes | Number of samples | Number of samples with 5-FU based chemotherapy | Number of OS events | Median follow-up time |
| --- | --- | --- | --- | --- | --- | --- |
| GSE39582 | Microarray | 20824 | 167 | 102 | 30 | 41.00 |
| GSE17538 | Microarray | 20824 | 232 | 88 | 40 | 47.83 |
| TCGA-COAD | RNA-Seq | 19462 | 456 | 122 | 18 | 25.15 |
| GSE106584 | Microarray | 23145 | 156 | 83 | 37 | 81.27 |
| GSE72970 | Microarray | 20824 | 124 | 124 | 92 | 22.80 |
| GSE87211 | Microarray | 20816 | 203 | 203 | 28 | 62.25 |

Consensus Weighted Gene Co-expression Network Analysis (WGCNA) is a powerful method that identifies clusters of genes (modules) with similar expression patterns across multiple datasets [34, 35]. This approach is particularly effective in integrating data from different sources [36], like tumor and organoid gene expression profiles in our study. By analyzing these patterns collectively on three datasets - colorectal tumors (GSE171680), colorectal tumor-matched organoids (GSE171681), and an independent colorectal organoid data (GSE64392), consensus WGCNA provides a more robust and comprehensive understanding of the underlying biological relationships. It helps in pinpointing key gene modules that are consistently associated across datasets, which are crucial for understanding complex traits such as drug response in cancer. The study then centers on hub genes within these modules, vital for developing a predictive model for chemotherapy response. This workflow combines matched tumor-organoid gene expression with additional drug response data, aiming to develop a robust predictive model for disease-relevant chemotherapy response.

To build and validate our drug-response prediction models, we incorporated a detailed methodological approach as depicted in Figure 1. Hub genes identified from the organoid WGCNA were employed to construct predictive models using ridge regression, random forest, and an ensemble of these methods. The models were trained and optimized through cross-validation to minimize prediction error, selecting the best-performing model based on its predictive accuracy in terms of area under the curve (AUC) on independent validation sets. The selected ensemble model demonstrated superior performance, indicating its effectiveness in predicting chemotherapy response. Further validation involved calculating patient-specific drug-resistance scores using the optimal model, allowing for a thorough assessment of individual response to chemotherapy.

### Identification of coherent gene modules through consensus WGCNA

Given the robust nature of the weighted correlation network to the choice of soft-thresholding power, we selected β = 12 for the signed network. This ensured a scale-free topology model fit above 0.75, in which the network conforms to a scale-free topology, a characteristic of biological networks where few nodes (genes) are highly connected [34]. (**Figure 2A**). The consensus WGCNA identified 16 modules, including a grey module. Grey modules typically contain genes that do not correlate well with any others and thus are not grouped into specific functional modules. In contrast, non-grey modules such as the turquoise module (n=214), blue module (n=190), brown module (n=157), yellow module (n=144), and green module (n=141), which contain the most genes, show more homogenous and potentially functionally relevant expression patterns. Although the grey module comprised 1910 genes, it displayed heterogeneous expression patterns and was not assigned to any particular function. **Figure 2B** shows the dendrogram of modules clustered by hierarchical clustering based on consensus Topological Overlap Matrix (TOM).

**Figure 2.**
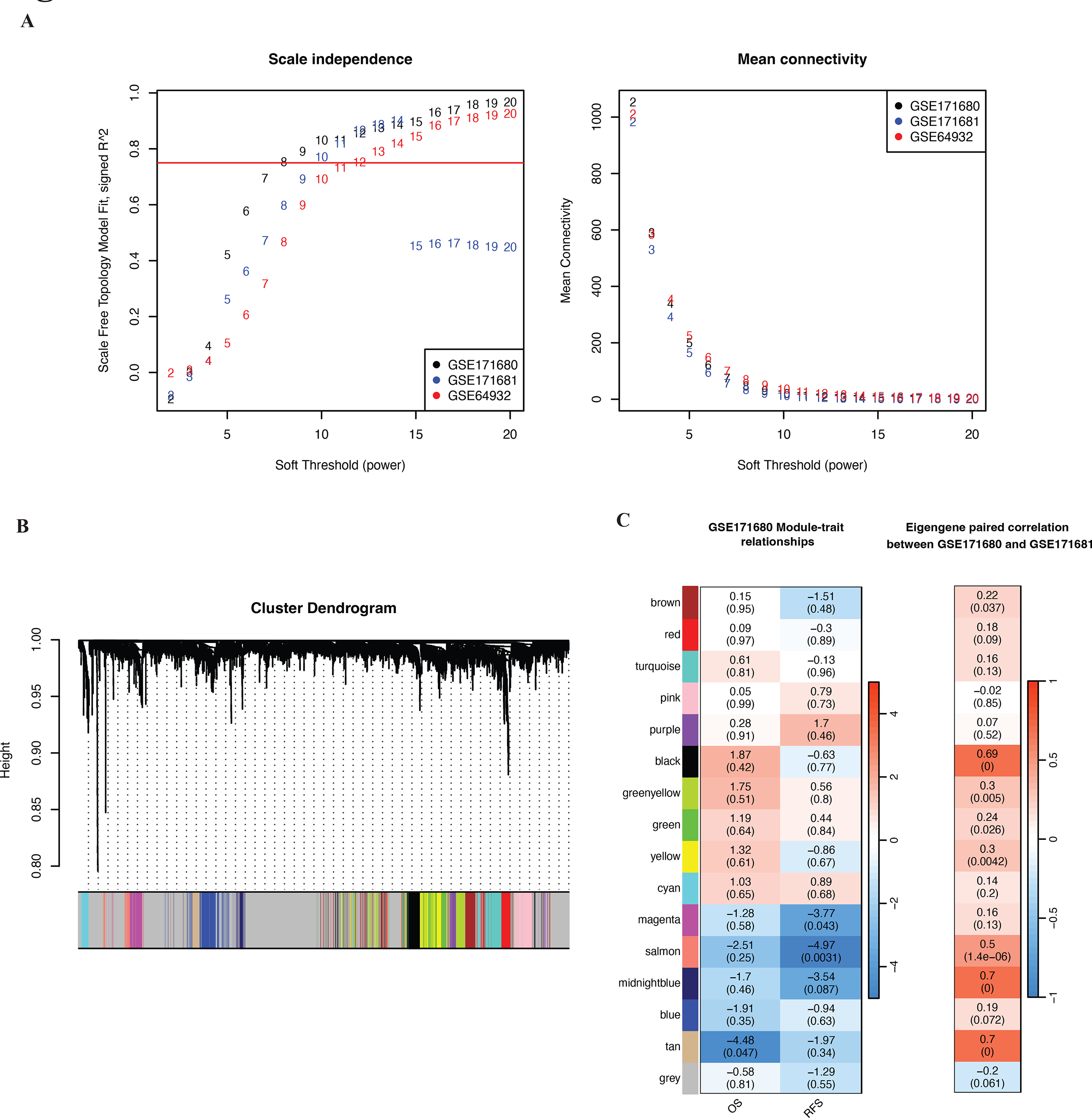
Construction of consensus WGCNA. (A) shows the scale-free topology model fit (y- axis) under different power values (x-axis); (B) shows the average connecWvity (y-axis) under different power values (x-axis); (C) shows the dendrogram of the consensus module clustering based on the dissimilarity measure (1- consensus TOM); (D) the heatmap on the lef plots the relaWonship of consensus module eigengenes and prognosis results of GSE171680; the heatmap on the right indicates the Spearman correlaWon between consensus module eigengenes of GSE171680 and GSE171681.

### Prognostic relevance and correlation of gene modules with clinical outcomes

Among the modules identified, the tan, salmon, and magenta modules showed statistically significant associations with Overall Survival (OS) and Recurrence-Free Survival (RFS) in Cox regression models in independent datasets (**Figure 2C**). These modules demonstrated protective effects, with hazard ratios less than one. Notably, the tan and salmon modules had significant Spearman correlations between their eigengenes in matched organoid (GSE171681) and tumor samples (GSE171682, suggesting robust biomarker potential. **Figure 3A** shows the scatterplot of Spearman correlations between the eigengenes of these two modules. The tan module, in particular, showed the highest correlation (*R*^2^_*tan*_ = 0.7, *p* = 2.2 × 10^%&’^ ), and the salmon module also achieves a moderate correlation coefficient (*R*^2^_*salmon*_ = 0.5, *p* = 1.4 × 10^%’^). **Figure 3B** shows the consensus network of the tan and salmon modules with the names of the hub genes colored in brown. The tan module has more connections than the salmon module. In each pair among the three datasets, the individual module membership Spearman correlations are all significant (**Figure 3C**). This indicates a high consistency of gene findings in the tan and salmon modules across the three datasets and confirms the reliability of these gene modules in predicting clinical outcomes. We identified and selected 35 hub genes with an absolute consensus module membership (MM) of 0.5 or higher for subsequent prediction modeling.

**Figure 3.**
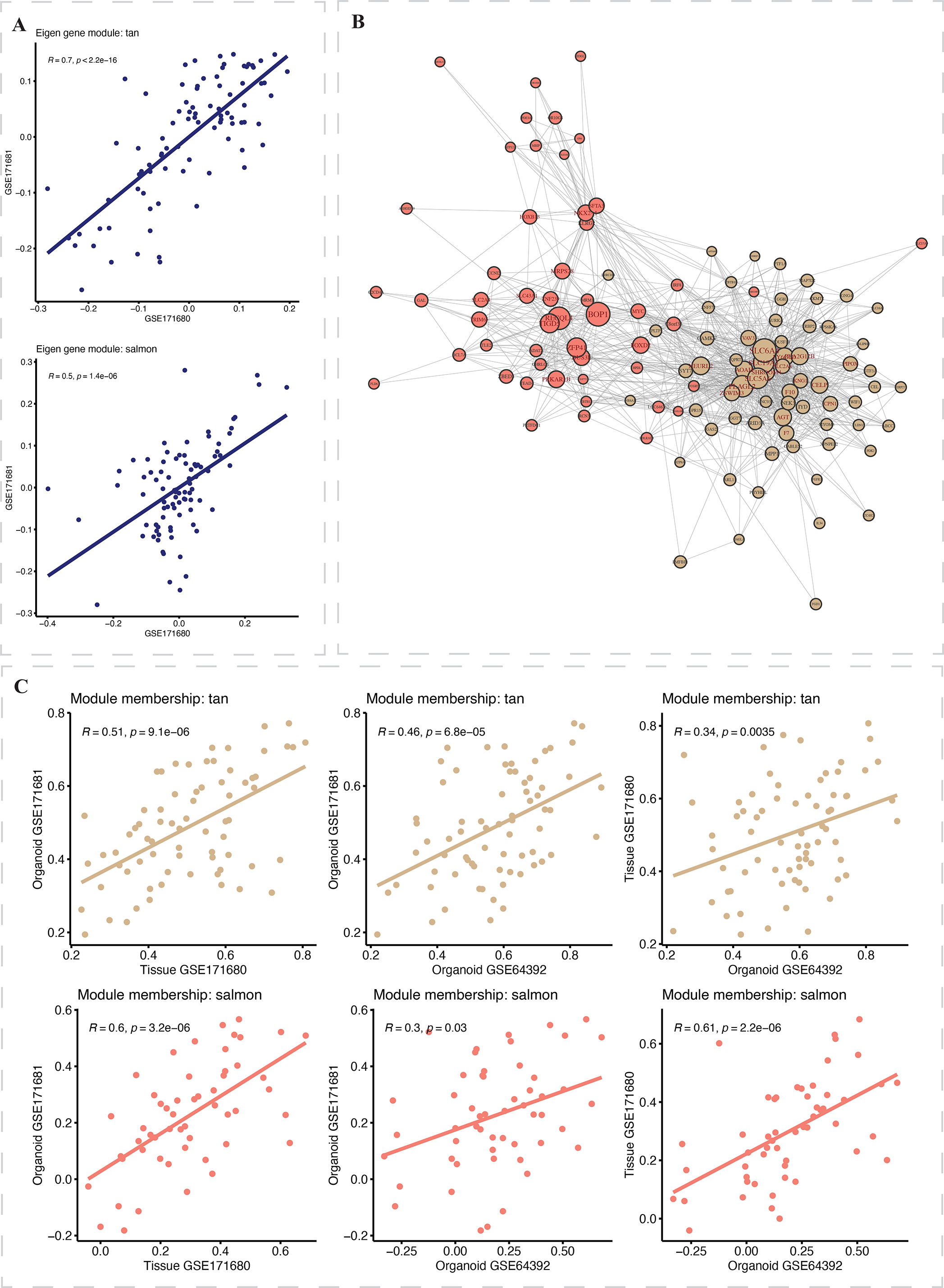
Plots of significant modules: tan and salmon. (A) shows the scaberplots between eigengenes of GSE171680 (x-axis) and GSE171681 (y-axis) of tan (top) and salmon (bobom), with Spearman correlaWons and P-values labeled; (B) shows the consensus network of the tan and salmon modules (hub genes colored in brown); (C) shows the scaberplots of the module membership (MM) among all three datasets of tan (top) and salmon (bobom) modules.

### Building organoid drug-response models and selecting drug response-related genes

The organoid 5-FU drug-response models were built using the 35 hub genes selected from WGCNA. To further identify the biomarkers following 5FU treatment in colorectal cancer, we developed an ensemble model that combined two machine learning methods: random forest and ridge regression (see “Methods”). We compared the performance of our ensemble method to that of using only ridge or only random forest methods through cross-validation in the training data GSE64392. We also validated these models on the GSE171680 dataset using the Area Under the Curve (AUC) metric (see **Supplementary** Figure 1). The results suggested that the biomarkers identified by the ensemble model demonstrated higher predictive performance across all models.

Using this model, seven genes were selected as the final biomarkers for the 5-FU drug response based on training data GSE64392 (**Table 2**).

**Table 2.** The seven genes selected by the ensemble organoid model.

| **Gene** | **Coefficient** | **Module** |
| --- | --- | --- |
| CELP | -0.0798 | tan |
| CPN1 | -0.0282 | tan |
| NEURL2 | -0.0162 | tan |
| PIPOX | -0.0392 | tan |
| SLC19A3 | -0.0568 | tan |
| VAV3 | 0.0012 | tan |
| HOXB13 | -0.0325 | salmon |

### Validation of organoid prediction model with six independent datasets

The prediction performance of our organoid model was then validated in six independent GSE datasets: GSE39582, GSE17538, GSE106584, GSE72970, and GSE87211. The details of the survival and transcriptomic data for these six datasets are described in **Table 1B**. The patient-specific drug-response scores of each validation dataset were calculated (see “Methods”) and the statistical difference in overall survival (OS) between the drug-sensitive and drug-resistant groups was assessed by the Kaplan-Meier survival curves and log-rank tests. As shown in **Figure 4** and **Table 3**, the drug-sensitive group had a significantly longer OS than the drug-resistant group for all six datasets: the p-values of log-rank tests are 1.32×10^-04^ (GSE39582), 7.09×10^-04^ (GSE17538), 4.95×10^-02^ (TCGA-COAD), 2.08×10^-02^ (GSE106584), 4.60×10^-03^ (GSE72970), and 1.25×10^-03^ (GSE87211). We examined the pattern of the seven selected drug-related genes.

**Figure 4.**
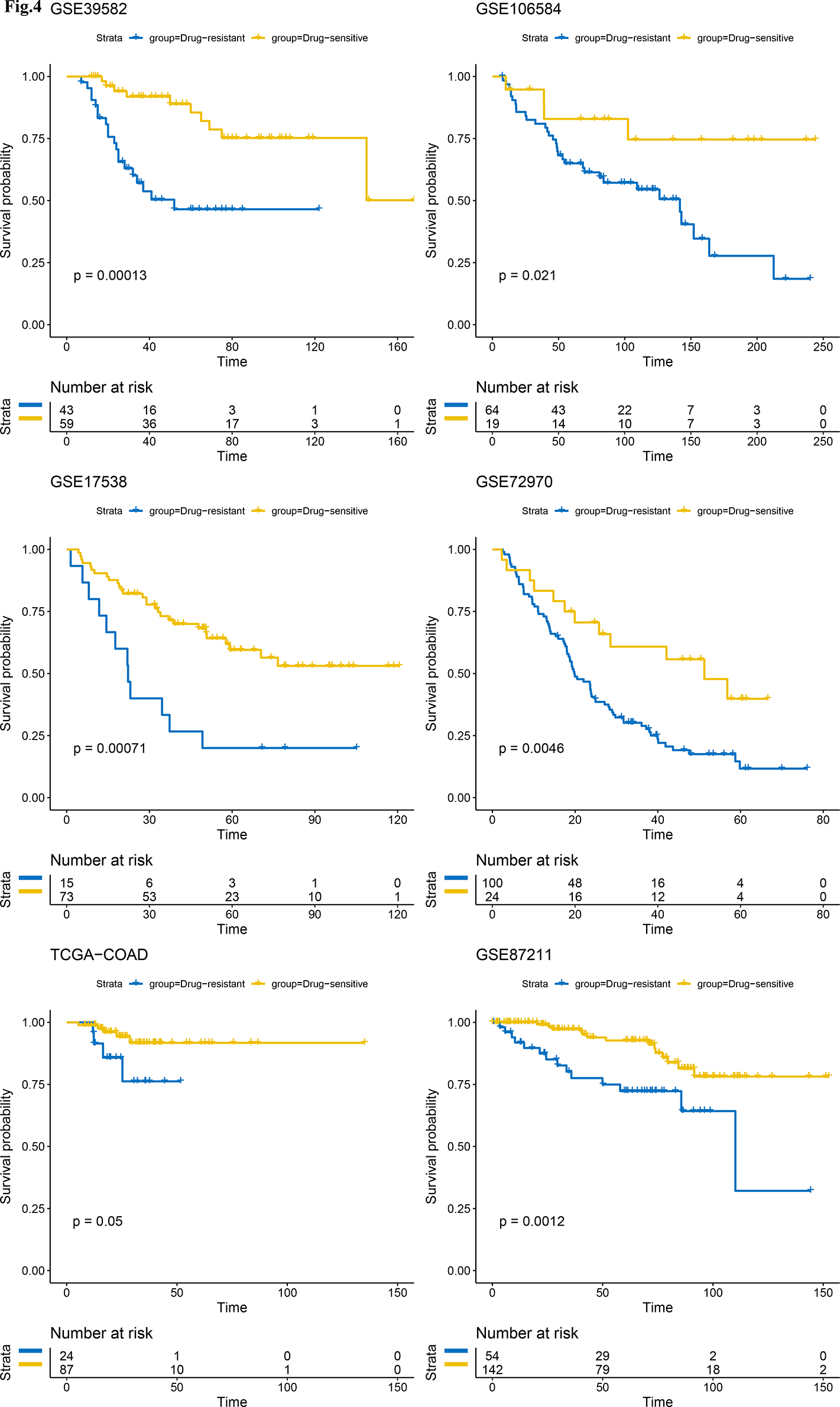
Drug-response predictions for 5-FU-based treated samples of six independent datasets. The predicted drug-resistant scores were divided into drug-sensiWve and drug-resistant group and tested on the overall survival results from six independent datasets. StaWsWcal significance was measured using Kaplan–Meier survival curves and log-rank tests. P-values <0.05 were considered significant.

**Table 3.**
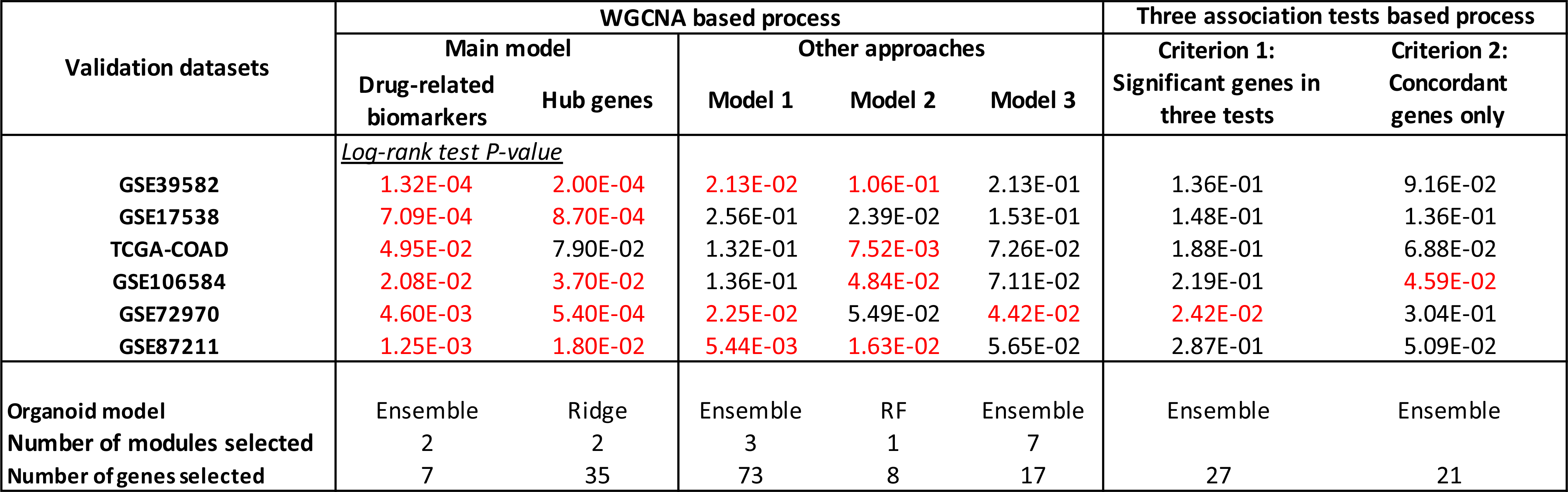
Prediction results of the main models and all other gene selection processes.

Interestingly, we found that all the genes with lower IC50 values show higher expression in GSE64932 (**Figure 5A**). Furthermore, to determine if the 35 hub genes identified in the consensus WGCNA have similar validation outcomes as the seven drug-related genes, we computed patient-specific drug-resistant scores using the ridge regression coefficients of the 35 hub genes. The prognosis test results are displayed in **Supplementary** Figure 2, which shows significant p-values were achieved in all datasets except TCGA-COAD, where the significance was marginally achieved. The selection of 35 hub genes from the WGCNA, and the further selection of 7 genes from these 35, provide a reliable predictor of drug response. This can be showed by the significant log-rank tests in 5 out of 6 validation datasets, which were based on survival outcomes of the 35 hub genes. Furthermore, all 6 validation datasets showed significant results in log-rank tests using the 7-gene organoid model.

**Figure 5.**
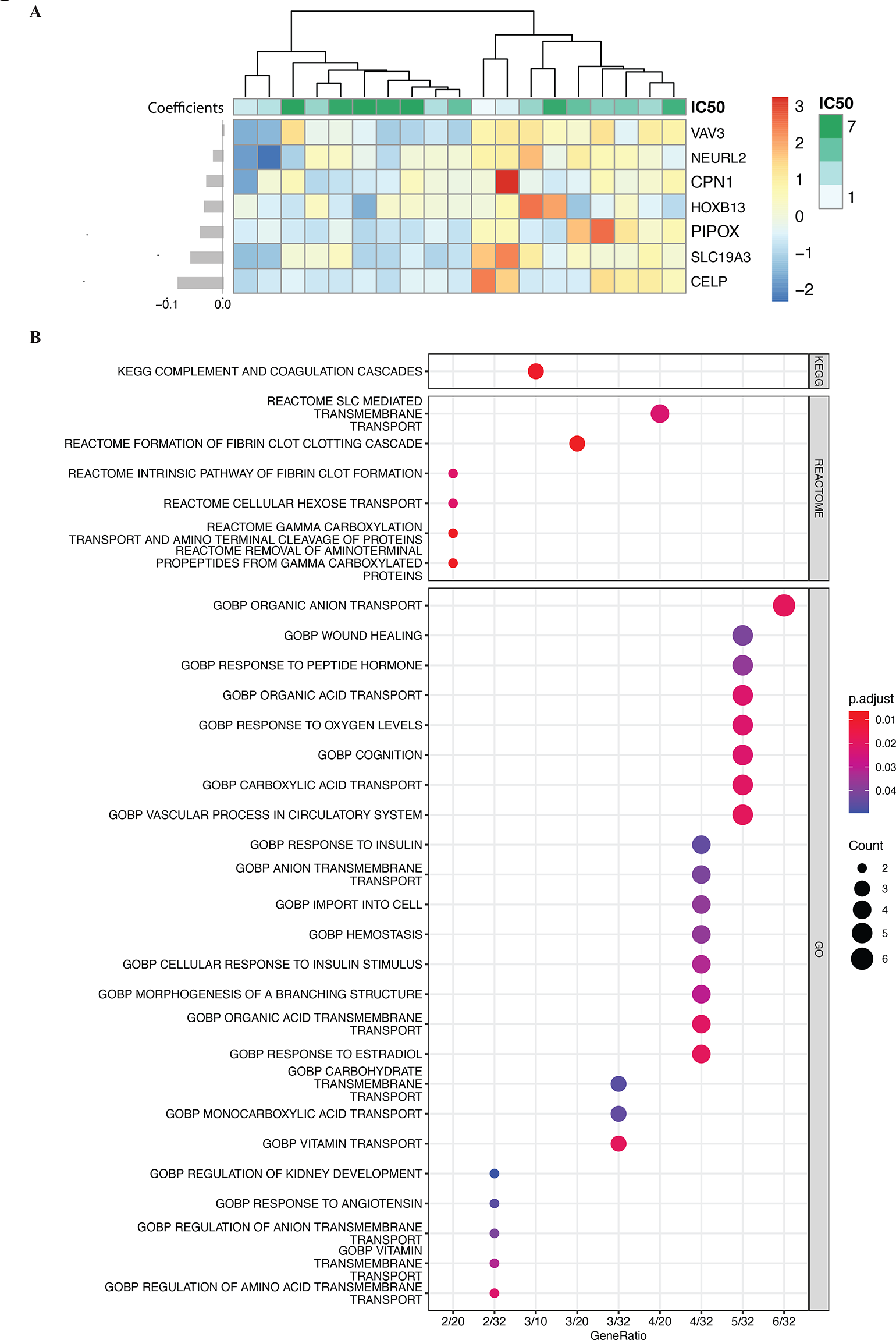
Heatmap of 7 drug-related biomarkers and functional enrichment plots of 35 hub genes selected from WGCNA. (A) displays a heatmap of seven drug-related biomarkers, accompanied by a hierarchical clustering dendrogram at the top. The coefficients of the biomarkers are depicted on the lef. (B) presents dot plots for the functinal enrichment analysis of 35 hub genes, selected from WGCNA. All significant pathways are included in the plot.

### Functional enrichment analysis of hub genes

We next performed enrichment analysis for the 35 selected hub genes in the KEGG, REACTOME, and GO pathway databases. **Figure 5B** presents the significant pathways with adjusted p-values less than 0.05. It can be observed that certain pathways are prominently associated with colon and rectal cancer chemotherapy response. Top pathways included those related to DNA repair mechanisms, cell cycle regulation, apoptosis, and drug metabolism. For instance, pathways involved in DNA damage response are crucial, as chemotherapy often targets rapidly dividing cancer cells by inducing DNA damage. Similarly, pathways regulating apoptosis is also significant, as the effectiveness of chemotherapy is partly determined by the ability of cancer cells to undergo programmed cell death.

### Prediction performance of alternative gene selection processes

To demonstrate the consistency and utility of our approach in predicting the survival of colorectal cancer patients, we evaluated the prediction performance on the validation datasets using two alternative gene selection methods: one based on three additional WGCNA strategies, and another based on two gene association tests (Table 3). We found that the genes chosen by Model 2 (See Methods), which were selected based on consensus WGCNA applied to two datasets, yielded significant results in the log-rank tests for four datasets (Supp Figure 4). This was followed by Model 1, where genes were selected based on WGCNA applied to tissue data GSE171680; it produced significant results in the log-rank tests for three datasets (Supp Figure 3). Model 3, in which genes were selected based on WGCNA applied to organoid data GSE64932, did not consistently yield significant results in log-rank tests (Supp Figure 5). This suggests that applying more data to construct consensus WGCNA yields more robust results in predicting survival. Next, we examined the gene selection process based on the two different criteria of gene filtering from the association tests. We found that the small sample size of the organoid and tissue data could potentially affect the reliability of test results, thereby impacting the validation results in survival prediction (Supp Figure 6-7).

## Discussion

In this study, we introduced a novel methodology for predicting chemotherapy responses in colon and rectal cancer, utilizing matched tumor-organoid gene expression data. Our approach incorporated Consensus Weighted Gene Co-expression Network Analysis (WGCNA) across diverse datasets, including colorectal tumors, corresponding organoids, and an independent CRC organoid dataset with drug response data (IC50 values). This method effectively identified key gene modules and hub genes associated with colorectal chemotherapy response, marking a significant advancement in personalized medicine approaches for colorectal treatment.

The prediction results of our study highlight the substantial advantages of using matched organoid and tumor samples for biomarker selection in colorectal cancer treatment. By integrating gene expression data from these matched samples, our analysis was able to identify more precise and relevant biomarkers for chemotherapy response prediction. This approach led to the discovery of specific gene modules and hub genes that are critically involved in response to chemotherapy in colorectal cancer, particularly 5-fluorouracil (5-FU). Our predictive model, built on these findings, demonstrated a notable improvement in accuracy and reliability compared to traditional methods as results showed in **Table 3**.

The superior performance of our proposed model underscores the effectiveness of the two strategies we employed. First, matched tumor-organoid data in capturing the intrinsic signature of chemo-response complex molecular dynamics of colon and rectal cancers, thus yielding a more robust clinical outcome. Second, the WGCNA network-based biomarker selection offered notable advantages in understanding colorectal cancer chemotherapy response. The ability of WGCNA to identify modules of co-expressed genes allowed us to discern complex gene interaction networks relevant to response to treatment in colon and rectal cancer. This network-based approach facilitated the identification of not just individual genes but also clusters of genes (modules) that collectively contribute to drug responsiveness. This method, by capturing the systemic relationships and dependencies among genes, provided a more holistic view of the molecular mechanisms underlying the response to chemotherapy in colorectal cancer, thereby enhancing the accuracy and relevance of the selected biomarkers for clinical application.

A limitation of the current work is the lack of direct drug response data from the matched colorectal cancer organoid dataset (GSE171681), necessitating reliance on similar in vitro organoid experiments (GSE64392). Future research could benefit from matched tumor-organoid datasets inclusive of patient and organoid drug response outcomes. Additionally, network modeling approaches could be refined to integrate more extensive biological information [37, 38], potentially offering more profound insights into CRC chemotherapy response mechanisms. The strength of this work highlights a new pathway in biomarker discovery for colon and rectal cancer chemotherapy response prediction, addressing typical high-dimensional genomic data challenges in cancer research like intratumoral heterogeneity and the curse of dimensionality. This method could extend to more advanced organoid cultures, including those that incorporate integrating the potential of organoid and the patient-tumor microenvironment [39], offering the potential to identify biomarkers for immunotherapy responses.

## Discussion

In this study, we introduced a novel methodology for predicting chemotherapy responses in colon and rectal cancer, utilizing matched tumor-organoid gene expression data. Our approach incorporated Consensus Weighted Gene Co-expression Network Analysis (WGCNA) across diverse datasets, including colorectal tumors, corresponding organoids, and an independent CRC organoid dataset with drug response data (IC50 values). This method effectively identified key gene modules and hub genes associated with colorectal chemotherapy response, marking a significant advancement in personalized medicine approaches for colorectal treatment.

The prediction results of our study highlight the substantial advantages of using matched organoid and tumor samples for biomarker selection in colorectal cancer treatment. By integrating gene expression data from these matched samples, our analysis was able to identify more precise and relevant biomarkers for chemotherapy response prediction. This approach led to the discovery of specific gene modules and hub genes that are critically involved in response to chemotherapy in colorectal cancer, particularly 5-fluorouracil (5-FU). Our predictive model, built on these findings, demonstrated a notable improvement in accuracy and reliability compared to traditional methods as results showed in **Table 3**.

The superior performance of our proposed model underscores the effectiveness of the two strategies we employed. First, matched tumor-organoid data in capturing the intrinsic signature of chemo-response complex molecular dynamics of colon and rectal cancers, thus yielding a more robust clinical outcome. Second, the WGCNA network-based biomarker selection offered notable advantages in understanding colorectal cancer chemotherapy response. The ability of WGCNA to identify modules of co-expressed genes allowed us to discern complex gene interaction networks relevant to response to treatment in colon and rectal cancer. This network-based approach facilitated the identification of not just individual genes but also clusters of genes (modules) that collectively contribute to drug responsiveness. This method, by capturing the systemic relationships and dependencies among genes, provided a more holistic view of the molecular mechanisms underlying response to chemotherapy in colorectal cancer, thereby enhancing the accuracy and relevance of the selected biomarkers for clinical application.

A limitation of the current work is the lack of direct drug response data from the matched colorectal cancer organoid dataset (GSE171681), necessitating reliance on similar in vitro organoid experiments (GSE64392). Future research could benefit from matched tumor-organoid datasets inclusive of patient and organoid drug response outcomes. Additionally, network modeling approaches could be refined to integrate more extensive biological information [37, 38], potentially offering more profound insights into CRC chemotherapy response mechanisms. The strength of this work highlights a new pathway in biomarker discovery for colon and rectal cancer chemotherapy response prediction, addressing typical high-dimensional genomic data challenges in cancer research like intratumoral heterogeneity and the curse of dimensionality.

This method could extend to more advanced organoid cultures, including those that incorporate integrating the potential of organoid and the patient-tumor microenvironment [39], offering the potential to identify biomarkers for immunotherapy responses.

## Method

### Study cohorts

We selected three colorectal cancer datasets from research carried out by van de Wetering et al.[22] and Cho et al. [28] for use in the consensus WGCNA algorithm and development of organoid drug-response model. The datasets from van de Wetering et al. contains 22 organoid samples of microarray data and drug-response for colorectal cancer. The microarray data can be retrieved from the Gene Expression Omnibus (GEO) with the study accession GSE64932 [22]. In our study, we used *IC*50 values as drug sensitivity measurements and selected 19 samples tested with 5-Fu. The two datasets from Cho et al. include paired expression data from 87 organoid (GEO accession: GSE171681) and patient tissue (GEO accession: GSE171680) samples with colorectal cancer. The detailed information of the three training datasets can be found in Table 1A. Furthermore, we validated the prognosis predictive value of our organoid drug-response model to the overall survival (OS) outcomes of five GEO datasets for colorectal and colon cancer (GSE39582 [40], GSE17538 [41], GSE106584 [42], GSE72970 [43], and GSE87211 [44]). We also included one TCGA data (https://www.cancer.gov/tcga) for colorectal cancer (TCGA-COAD). All six validation datasets contain OS outcomes and the samples with 5-FU based treatment were selected. The detailed information of validation data can be found in Table 1B.

### Data preprocessing

For the expression datasets from GEO, we used the robust multichip average (RMA) normalized expression data [45]. The genes were represented by the probes with the largest interquartile range (IQR) statistics using *findLargest* function in genefilter R/Bioconductor package. The gene symbols were annotated using the AnnotationDbi R/Bioconductor package. The TCGA-COAD patient data were downloaded from the TCGA data portal using the TCGAbiolinks Bioconductor/R package [46]. For expression analysis of TCGA-COAD, we used the FPKM-UQ dataset and performed a log2 transformation.

### Consensus WGCNA

We used consensus WGCNA to study the relationships among the three expression profiles (GSE64932, GSE171680, and GSE171681) [34]. To perform this analysis, we first filtered the three expression profiles, selecting only the 3637 common genes that were among the top 50% most variable genes across all datasets. For each dataset, a signed correlation weight, *S*_*ij*_ = 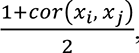 , is assigned to each gene pair *x*_*i*_ *and x*_*j*_ via a positive soft thresholding parameter *β*.

The signed network weighted adjacency matrix is defined as:

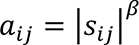

Here, *β* is the raised power of similarity measures, which emphasize more on the strong associations [47]. The three adjacency matrices were then transformed into the topological overlap matrices (TOM) that provide a robust measure of connections between gene pairs [48]. The individual TOMs are calibrated using the full quantile normalization such that all quantiles equal each other. To obtain the consensus TOM, we calculated the component-wise mean of individual TOMs for each set. The genes were further clustered by average linkage hierarchical clustering using the dissimilarity measure of the consensus TOM (1 – consensus TOM). The dendrogram cut height for module detection was set to 0.999 and the minimum module size of each module was set to 30 genes. The consensus network analysis was conducted using the *blockwiseConsensusModules* R function in WGCNA package.

### Significant modules and hub genes selection

To identify the significant modules, we first performed Cox proportional hazards regression models to test association between patient OS and recurrent free survival (RFS) outcomes with the module eigengenes (MEs). This approach was adopted as the survival outcomes are significantly related to drug response. The module eigengenes were obtained as the 1_(!_ principal component of the patient tissue expression. Three modules were selected with coefficient *P-value* < 0.05 in either OS or RFS cox regression model. Furthermore, to determine the concordance of modules between organoid and tissue expression, we estimated Spearman correlations between the eigengenes of each module in the two paired organoid and tissue expressions (GSE171681 and GSE171682). Only two modules, tan and salmon, among the three survival outcomes significant modules, were highly significant correlated (*R*^2^*_tan_* = 0.7, *p* = 2.2 × 10^-16’^ and *R*^2^*_salmon_* = 0.5, *p* = 1.4 × 10^-16^) between their eigengenes. These two modules were then selected as the significant modules for further model training. To assess the module membership (MM) of genes in each module, we calculated the correlation between the gene expressions and MEs of each module. This is denoted as kME. To evaluate the MM across all the expressions, we used the consensus kME that obtained by average aggregation of the kMEs for each expression set. The consensus kME was implemented in the function *consensusKME*. The selection of hub genes varies as each dataset has different clinically related information. For example, GSE64932 only contains drug response data, while GSE171681 only includes survival outcome data. We selected the hub genes of the significant modules using only the criterion of consensus |MM| ≥ 0.5.

### Organoid drug-response model training

To build the drug-response models, we used the hub genes selected from the WGCNA of the organoid expression profile against the median IC50 of 5-FU as drug response. We selected ridge, random forest, and an ensemble method of random forest and ridge as the training models. Elastic net model was performed using the glmnet R package. The optimal *α* and *λ* parameters were selected based on the lowest mean squared error between the drug response and predicted values in the validation sets, using 3-fold cross validation (CV). This was implemented in the *cv.glmnet* function. The Random Forest (RF) was constructed using the *rfsrc* function from the randomForestSRC R package. The final RF model was built using genes with a permutation importance greater than 0. The ensemble method was created by choosing genes based on their permutation importance obtained from random forest model. These selected genes were then fitted into a ridge regression model. Following the selection of drug-response related genes, we conducted a 3-fold CV on each model to decide the optimal model as our final organoid model. Furthermore, to ensure the optimal model is selected, we applied the three models to patient data GSE171680, which was used to conduct the consensus WGCNA. The Area Under the Curve (AUC) was calculated based on the predicted values of each model, using the OS outcome as binary. The ensemble model was selected as the optimal model as it achieved both lowest 3-fold CV values and highest AUC among all three model. To stabilize the CV errors and assess the model powers, we repeated running the 3-fold CV for each model with 100 times. Seven genes were selected by the optimal model as the drug-response related genes.

### Patient specific drug-resistant score

To validate our optimized organoid drug-response model, we calculated the patient specific drug-resistance score for each patient using the corresponding expression data. Specifically, the score from the optimized organoid drug-response is calculated as follow:

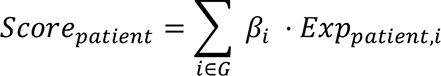

where *i* is gene from the 7 drug-response related genes *G, Exp*_*patient,i*_ represents the expression level of gene *i* of the patient, and *β*, is the ridge regression coefficient of gene *i* from the optimized organoid model. In the random forest model, the patient-specific score is derived from the predicted values. These values are estimated by the model using the expression data of the selected genes. For each validation dataset, the drug-resistant scores were separated into two groups: the drug-resistant group (with score ≥ cut point) and the drug-sensitive group (with score < cut point). The maximum rank statistic, which implemented in the MaxStat R package, was used to determine the cut point for each validation dataset. The Kaplan-Meier survival analysis and the log-rank test were used to visualize and evaluate the statistical differences in overall survival (OS) between the two groups.

### Functional enrichment analysis of hub genes

The pathway analysis was performed for the selected 35 hub genes using over representation analysis, which was implemented in clusterProfiler R package [49]. Briefly, this method determined whether biological processes that were over-represented in the gene list of interest using p-values calculated by hypergeometric distribution and adjusted by Benjamini-Hochberg (BH) method to calculate false discover rate (FDR). For the selected hub genes, we analyzed pathways from KEGG, Reactome and GO pathways from the Molecular Signatures Database (MSigDB), which can be accessed by the msigdbr R/Bioconductor package. The minimum and maximum sizes of gene sets used for analysis were set to 10 and 500 respectively.

### Gene selection process based on other WGCNA approaches

Additionally, we performed three other WGCNA models to select candidate genes: Model 1, WGCNA based on only the tissue expression (GSE171680); Model 2, consensus WGCNA based on the matched organoid and tissue expressions (GSE171681 and GSE171680); and Model 3, WGCNA based on only the organoid expression with drug response (GSE64932). For Model 1 and 2, the modules were identified as significant based on the Cox regression results regarding the OS outcome. We filtered the hub genes for further model training using |MM| ≥0.5. As there was no public drug response data available for this study, we used the OS outcome as response to train three models: ridge, random forest, and the ensemble model. On the other hand, the significance of the modules of Model 3 was determined by the Spearman correlation between the *IC*_50_ and the eigengene of each module. The same criterion was conducted to select the hub genes and followed by training the organoid models.

### Gene selection process based on gene association tests

We compared gene selection methods by conducting gene filtering based on the results of three association tests: 1) Gene-OS Cox regression test, 2) Gene-drug response Spearman correlation test, and 3) Gene-paired Spearman correlation test. Specifically, for the gene-OS Cox regression test, we fit the model to the OS outcome of GSE171680, with each gene as the dependent variable. Each model was adjusted for age and sex to account for confounding variables. The gene-drug Spearman correlation test was conducted between the *IC*_50_ drug response and each gene of GSE64932. The gene-paired Spearman correlation test was conducted between the gene expression levels of the paired organoid and tissue data. The candidate genes of training the organoid models were selected based on their significance in gene-paired correlations, gene-OS association tests and gene-drug correlation tests. We employed two gene filtering criteria and built organoid models using the genes chosen based on these criteria respectively. The first criterion selects genes where all test result p-values are smaller than 0.05. The second criterion selects genes where all test result p-values are smaller than 0.05 and the sign of the gene-OS and gene-drug coefficients are in agreement.

## Data Availability

All datasets used in the study are publicly available data from Gene Expression Omnibus (https://www.ncbi.nlm.nih.gov/geo/) and NCI Genomic Data Commons Data Portal (https://portal.gdc.cancer.gov/)

https://portal.gdc.cancer.gov/

---

**Supplementary** Figure 1 **- Showcases the cross-valida/on (CV) errors and ROC curves of the Ridge, RF, and ensemble models.** On the lef are boxplots of 100 repeated CV errors. The t-test results between Ridge vs ensemble and RF vs ensemble are displayed at the top of the boxplots, where “**” represents P-values < 0.01 and “****” signifies P-values < 0.0001. On the right are the ROC curves from tesWng on the binary label of the OS results of GSE171680 across the three models. The area under the curve (AUC) values are labeled for each model.

**Supplementary** Figure 2 **- Drug-response predictions for 5-FU-based treated samples of six independent datasets with 35 hub genes selected from the consensus WGCNA**

**Supplementary** Figure 3 **- Drug-response predictions for 5-FU-based treated samples of six independent datasets with candidate genes selected from WGCNA Model 1**

**Supplementary** Figure 4 **- Drug-response predictions for 5-FU-based treated samples of six independent datasets with candidate genes selected from WGCNA Model 2**

**Supplementary** Figure 5 **- Drug-response predictions for 5-FU-based treated samples of six independent datasets with candidate genes selected from WGCNA Model 3**

**Supplementary** Figure 6 **- Drug-response predictions for 5-FU-based treated samples of six independent datasets with candidate genes selected from filtering criterion 1 of gene associa/on tests**

**Supplementary** Figure 7 **- Drug-response predictions for 5-FU-based treated samples of six independent datasets with candidate genes selected from filtering criterion 2 of gene associa/on tests**

## DECLARATIONS

### Ethics approval and consent to participate

Not Applicable

### Availability of data and materials

All datasets analyzed in this study are publicly available and listed in Table 1. The analysis code is available at https://github.com/TransBioInfoLab/Organoid-Prediction.

### Competing Interest

The authors declare that they have no conflicts of interest.

### Funding

This research was supported by US National Cancer Institute grant R37CA248289 (J.J.S), and Sylvester Comprehensive Cancer Center Intramural program SCCC-NIH-2022-11 (X.S.C).

## Notes

### Competing Interest Statement

The authors have declared no competing interest.

### Summary of Updates

The abstract and Results section were updated; Figures 1 and 2 were revised

