## Supplementary figures and images for "Enhancing Chemotherapy Response Prediction via Matched Colorectal Tumor-Organoid Gene Expression Analysis and Network-Based Biomarker Selection"

### Supplemental Figure 1 to Figure 7

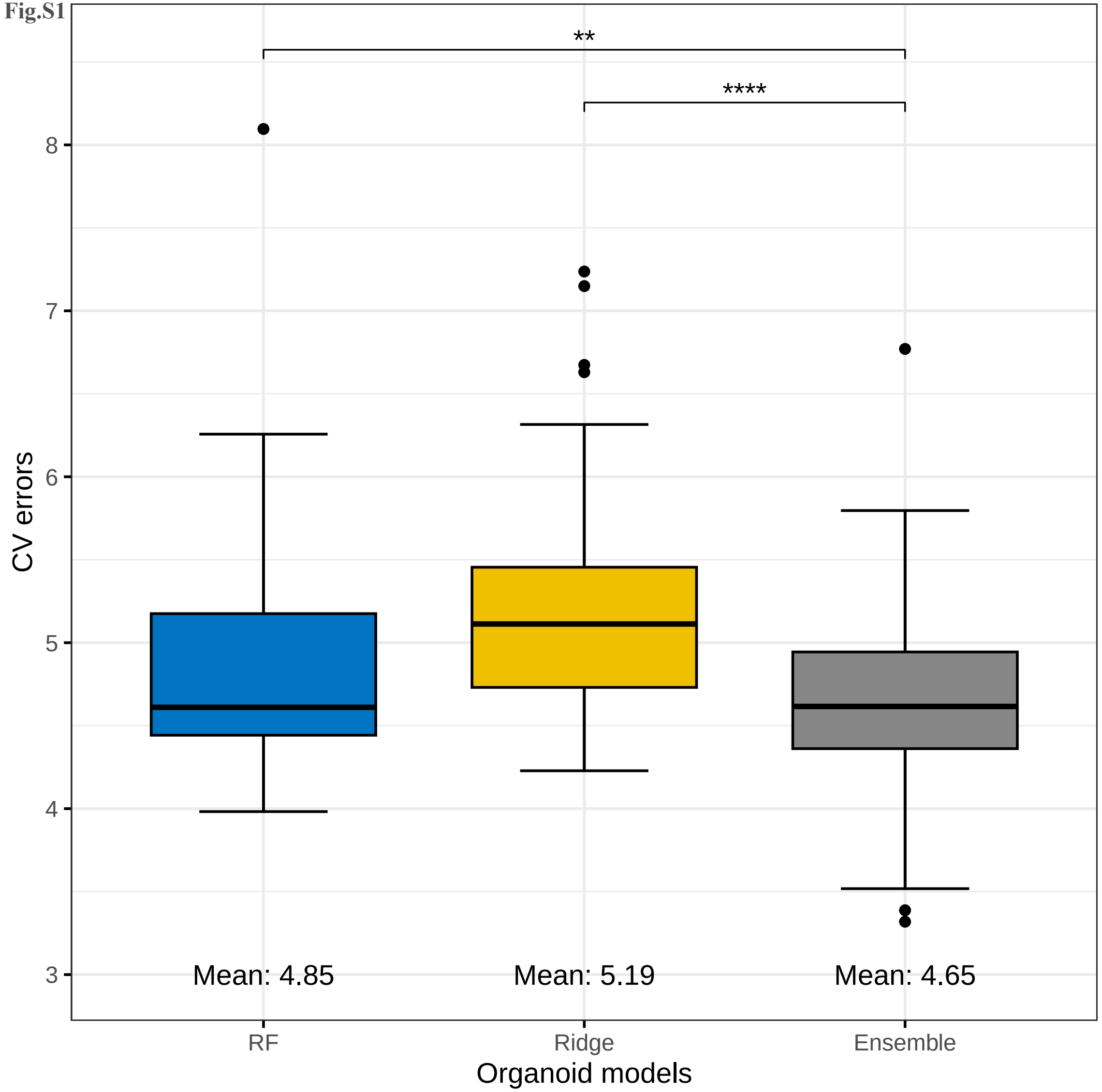

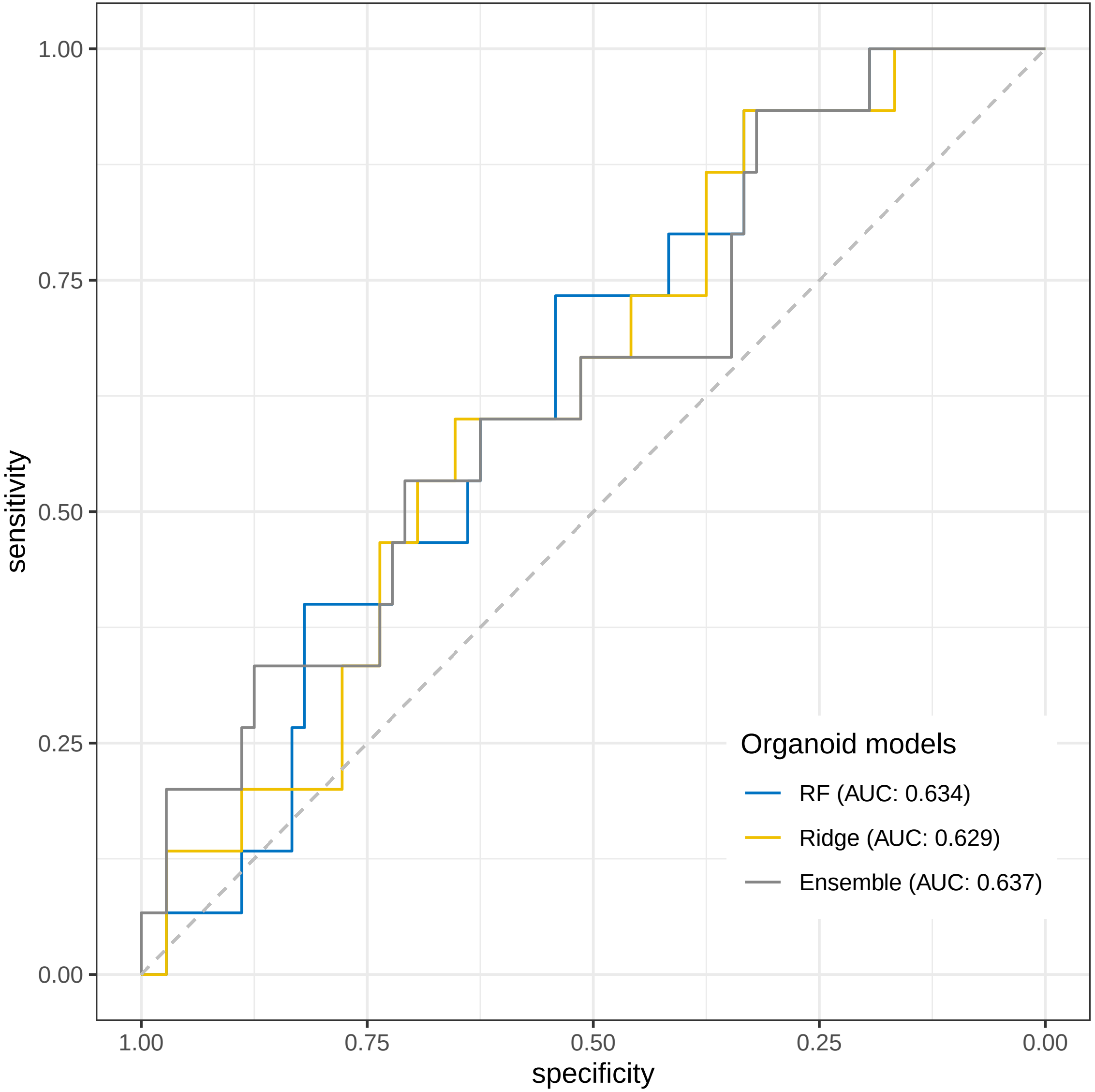

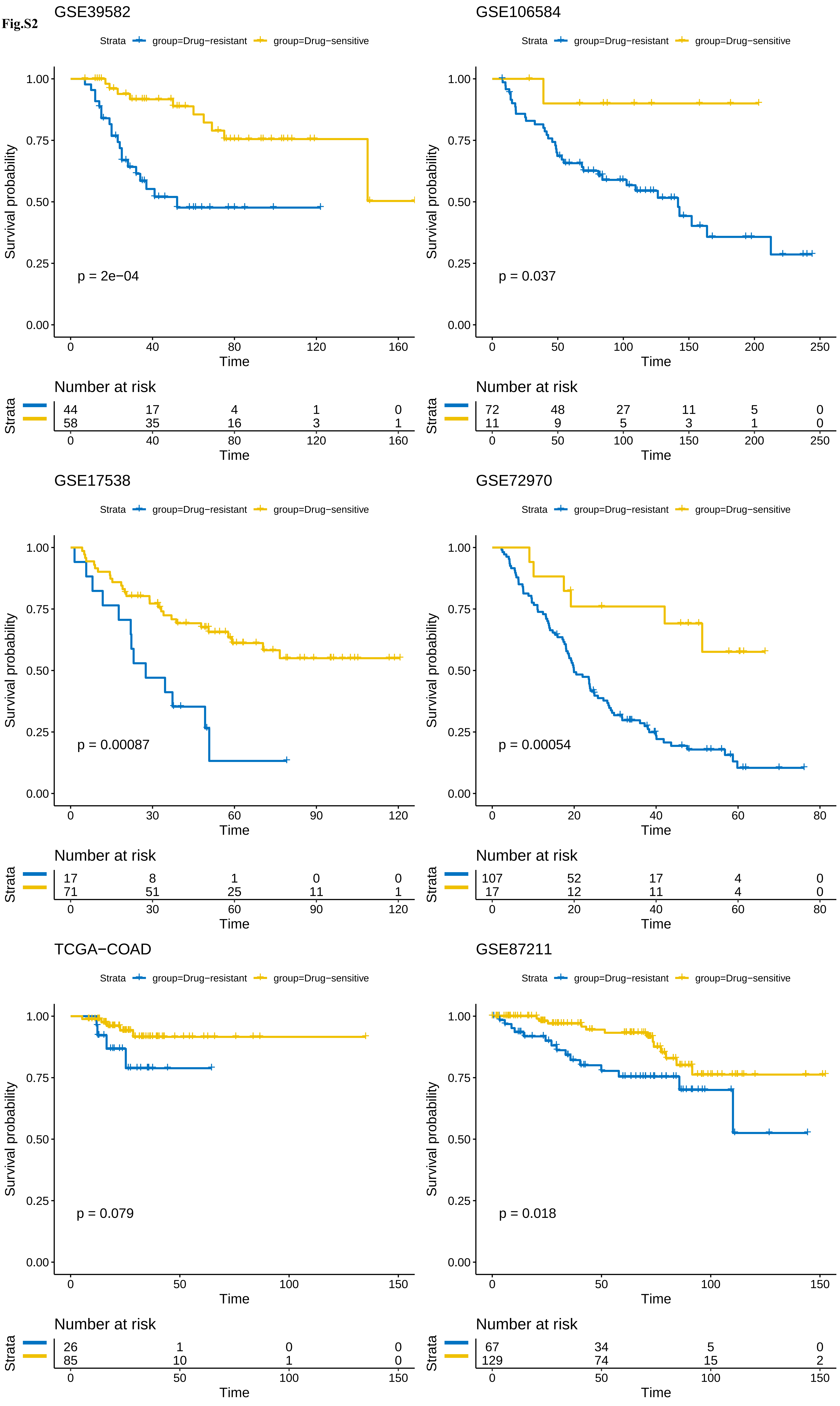

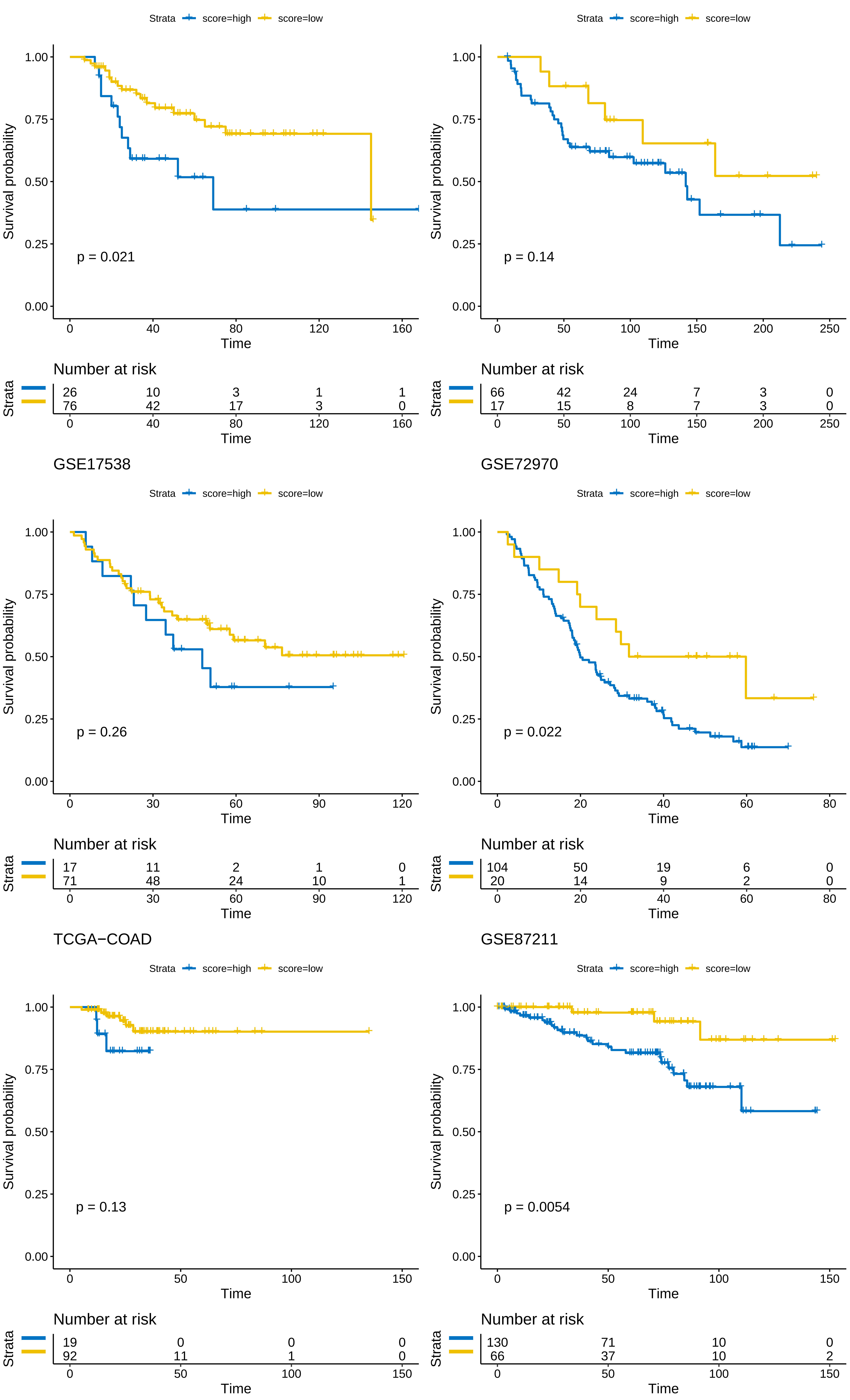

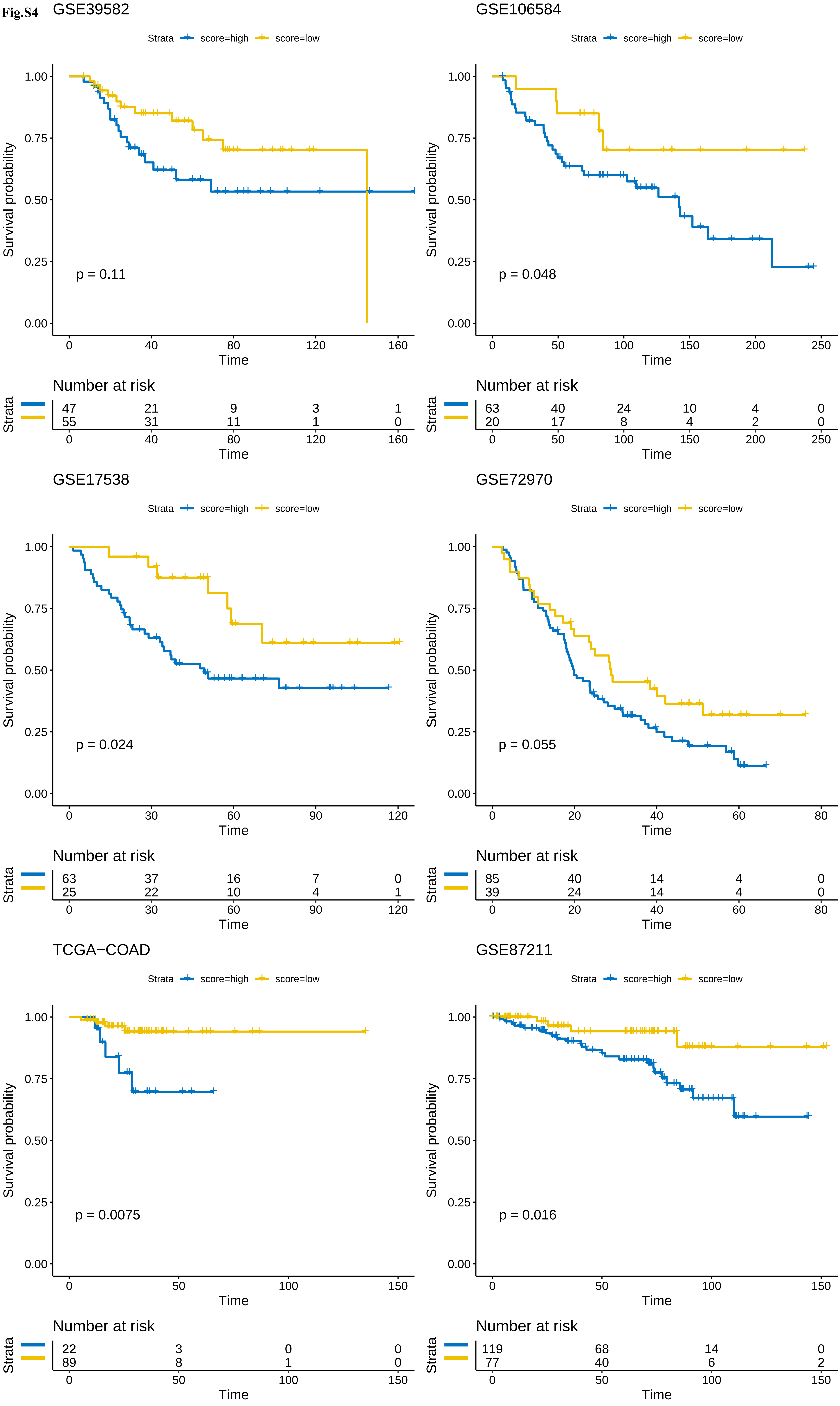

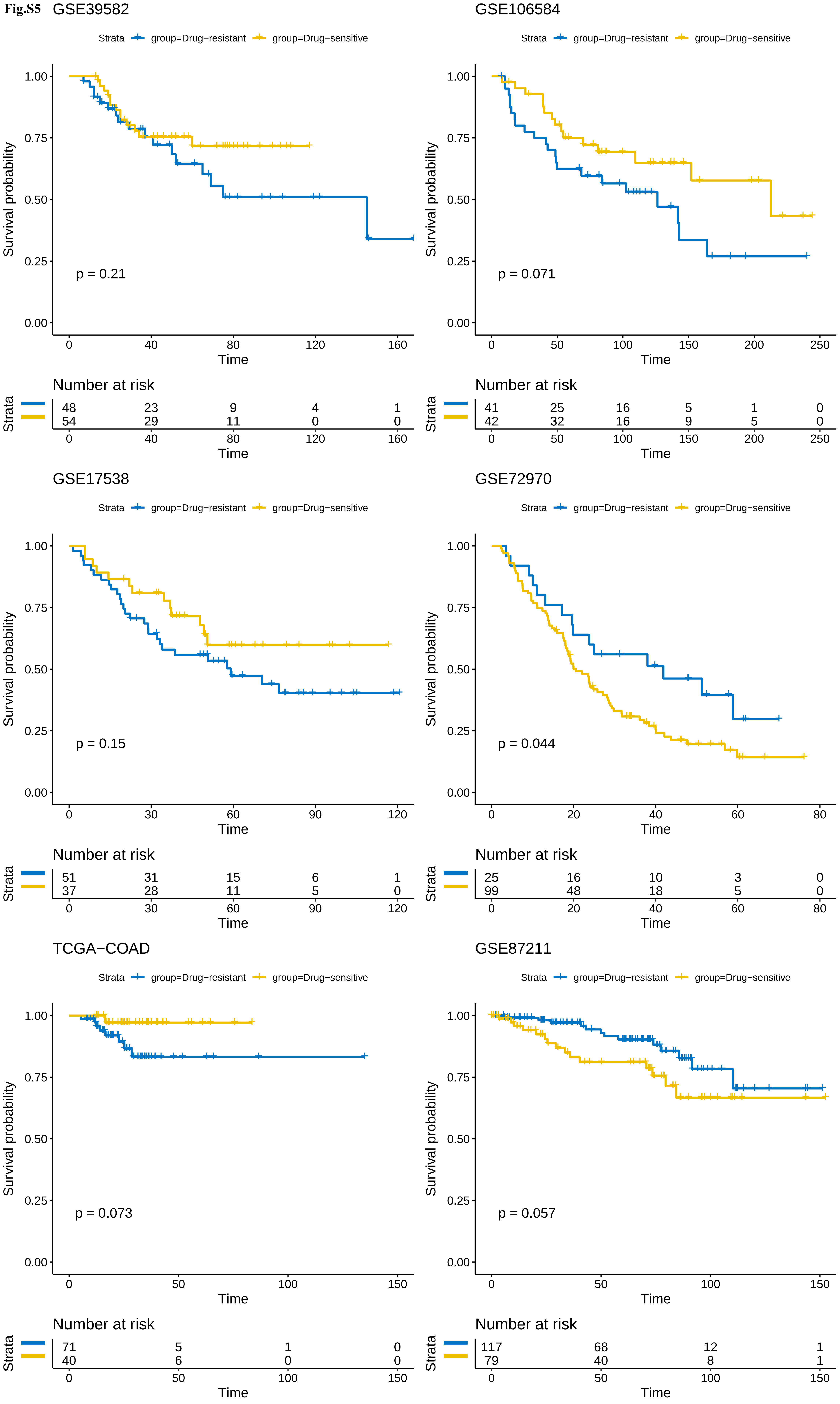

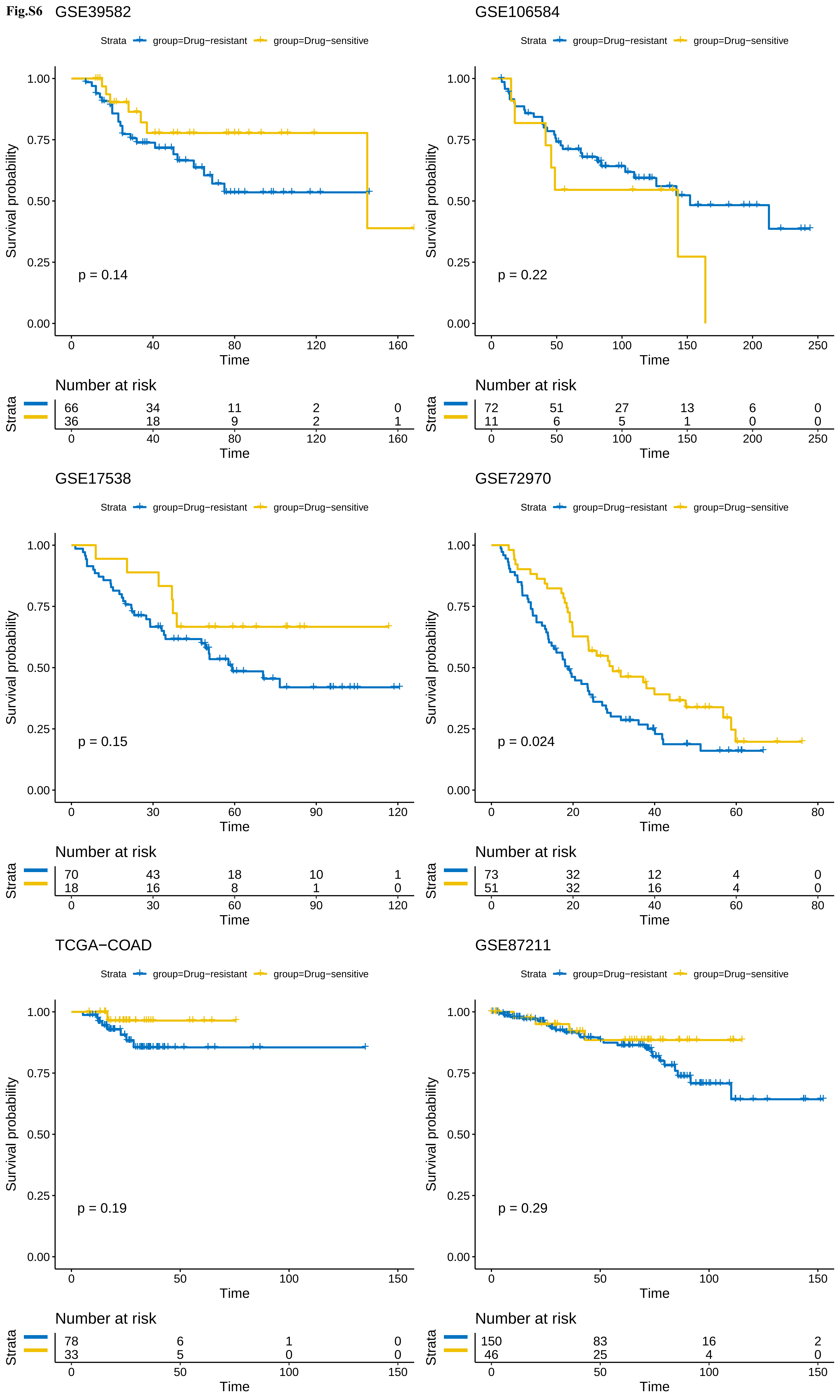

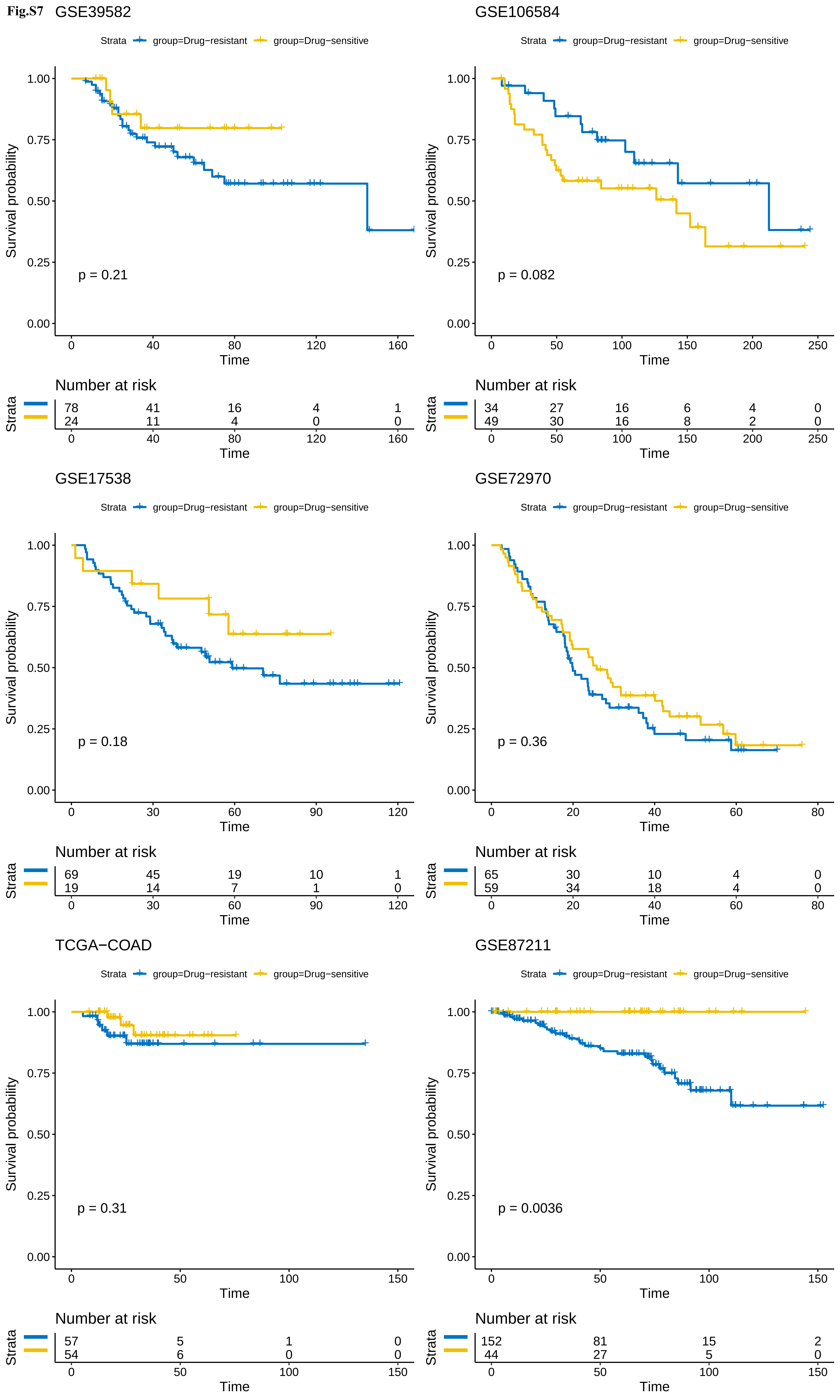
